# Application of tailored motor learning in community-based physiotherapy: a user-centered design and process evaluation of an extended framework

**DOI:** 10.1101/2024.06.07.24308412

**Authors:** Guus Rothuizen, Li-Juan Jie, Gaston Jamin, Roderick Wondergem, Susy Braun, Melanie Kleynen

**Affiliations:** Research Center for Nutrition, Lifestyle and Exercise, School of Physiotherapy, Zuyd University of Applied Sciences, Faculty of Health, Heerlen, The Netherlands; Department of Health Innovations and Technology, Fontys University of Applied Sciences, Eindhoven, The Netherlands

**Keywords:** geriatric individuals, neurological disorders, physiotherapy, motor learning

## Abstract

**Objective:** To develop supplementary knowledge and tools to support the application of motor learning in community-based physiotherapy of geriatric individuals and evaluate physiotherapists’ experiences of the developed knowledge and tools.

**Design:** A prospective case study comprised of two phases: 1) a user-centered design, and 2) a process evaluation.

**Setting:** Community-based physiotherapy practices.

**Participants:** Five physiotherapists were included for the user-centered design and another eight for the process evaluation. Making a total of thirteen participating physiotherapists during this study.

**Intervention:** Not applicable.

**Main outcome measures:** To evaluate the use of the physiotherapists with the extended framework a biweekly monitor was administered. To evaluate the experience of the physiotherapists with the extended framework three digital questionnaires were administered, and a midterm and final in-person evaluation were organized.

**Results:** The user-centered design resulted in a card deck and website with different layers of practical examples and theoretical information. Most of the participating physiotherapists (n = 13) barely used the extended framework during the evaluation period. Commonly reported reasons for not using the extended framework included a lack of time as well as the material’s not fitting into the physiotherapists’ daily routines. They reported, however, that the extended framework increased their motor-learning knowledge and confidence. Some motor-learning strategies were applied much more frequently than others in daily practice. The underlying reasoning regarding the application of some strategies over others varied widely.

**Conclusion:** The results indicate that physiotherapists felt unambiguous regarding the extended framework. The time and energy cost to breaking one’s own routines might have outweighed the potential benefits. Future research should aim to determine whether the extended framework applies similarly in different settings.

---

Many geriatric individuals with or without a neurological disorder (e.g., dementia, Parkinson’s disease, and osteoarthritis) experience difficulties when performing daily activities, such as walking, cycling, dressing, and performing chores around the house.^1,2^ The ability to move safely and efficiently is also essential for maintaining an active lifestyle and preventing new complaints and injuries.^3^ Physiotherapists specialize in supporting patients in the process of learning and improving movements, referred to as *motor learning* and reflects a relatively permanent change in a person’s capability to perform a motor skill.^4^

In recent years, the motor-learning field has experienced substantial growth, presenting physiotherapists with the challenge of effectively navigating and incorporating the expanding body of knowledge.^5,6^ In addition, since pressure on the healthcare system is increasing in the West due to the aging population, time is increasingly scarce for physiotherapists.^7,8^ In addition to the amount of literature, the content of the research also poses a challenge. Namely, various motor-learning strategies have been theoretically described and studied in a laboratory setting but lack guidance regarding application in clinical practice.^9,10^ In addition, research has stressed that individual factors, such as cognitive abilities, level of impairment, and personal preferences, should determine the motor-learning approach.^11–14^ Deciding which motor-learning approach optimally matches the patient seems to require substantial scientific knowledge, experience, and creativity from the therapist.^15^ It is, accordingly, not surprising that therapists struggle to apply motor-learning theory in practice. The problem of transferring theory into practice is called the *knowledge-to-practice gap* and is considered a key problem in the field of motor learning.^5,16^

To support clinicians’ decision-making and aid evidence-based implementation of motor-learning strategies in their clinical practice, Kleynen and colleagues developed a practical framework based on the broad distinction between conscious and non-conscious attributes of the motor-learning process.^15^ The distinction proposes that implicit motor learning targets more unconscious attributes of the motor-learning process, whereas explicit motor learning targets more conscious attributes.^17^ The framework includes seven common motor-learning strategies that have been categorized as promoting more implicit or explicit motor learning: errorless learning, dual-task learning, analogy learning, discovery learning, observational learning, movement imagery, and trial and error learning. The framework was informed by practice-based evidence of experts from different fields (e.g., researchers, healthcare professionals), as well as by research results that underpin the working mechanisms of these different learning strategies. Still, while it conveys structure to the theoretical foundation of motor learning, clear practical guidance remains limited. This is particularly evident for community-based physiotherapists, who encounter a diverse patient population in terms of age, disability, cognitive ability, and preferences, placing significant stress on their clinical decision-making process.^18^ It seems necessary to further expand and substantiate the framework with practical knowledge and tools to assist physiotherapists in the application of motor learning.^15^

This study, therefore, addressed the following research questions: 1) Which supplementary knowledge and tools (extensions) should be developed to support the application of the framework for motor learning in geriatric individuals (with or without a neurological disorder) in community-based physiotherapy practices? 2) How do physiotherapists use and experience the extended motor-learning framework in community-based physiotherapy practices?

## Study design in two phases

This prospective case study comprised two phases: the user-centered design of the extended framework and a process evaluation consisting of six months’ application of the extended framework (*Fig. 1*). The user-centered design (Phase 1) was described according to the four subphases, while the process evaluation (Phase 2) was divided into a methods and a results section. The study protocol (reference number METCZ20230061) was approved by the Ethics Committee from Zuyderland Hospital, Heerlen, The Netherlands.

In total, 10 community-based physiotherapy practices were involved in the study. Physiotherapists working in these practices were approached and screened for eligibility. The physiotherapists were eligible for inclusion when they were working in a community-based practice, had at least one year of working experience, and were treating older adults (65 years or older) with or without a neurological disorder. Five practices participated in the user-centered design. The process evaluation included another five practices that had not been involved in the design process. Participants signed an informed-consent form prior to joining the study.

**Figure 1:**
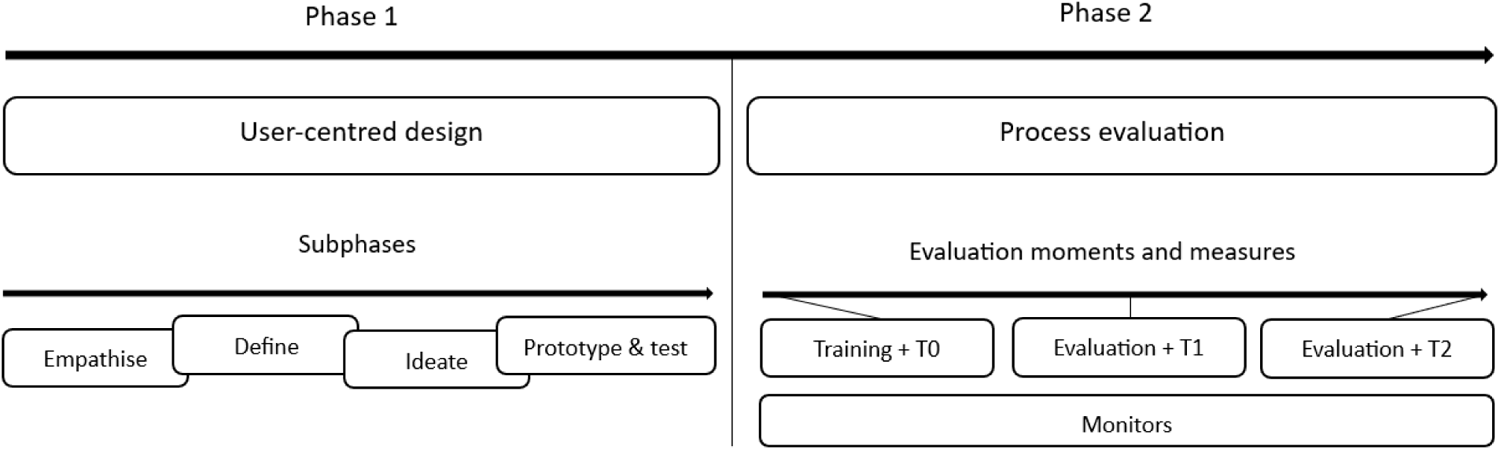
Study overview in two phases. The first phase focused on the development of an extended framework with a user-centered design. In the second phase, the framework with supplementary knowledge and tools was evaluated. Training = the preparatory training, evaluation = the in-person evaluation meetings, T0 = the digital questionnaire at baseline, T1 = the digital questionnaire at three months, T2 = the digital questionnaire at six months.

## Phase 1: User-centered design of the extended framework

The user-centered design contained four phases: empathize, define, ideate, and prototype and test^19,20^ and was conducted with five self-selected physiotherapists from five different community-based practices. A relatively small number of participants was previously shown to be sufficient for the design process.^21^ The therapists ranged from 30 to 56 years of age and had between eight and 33 years of work experience.

The *empathize phase* focused on understanding the context of users.^22^ Two in-person sessions were planned to explore the challenges physiotherapists face in daily practice and their needs regarding the application of motor learning. Subsequently, these challenges and needs were specified by the research team in the *define phase* and processed into fictional personae (*Fig. 2*) that represented the participating physiotherapists.^23^

**Figure 2:**
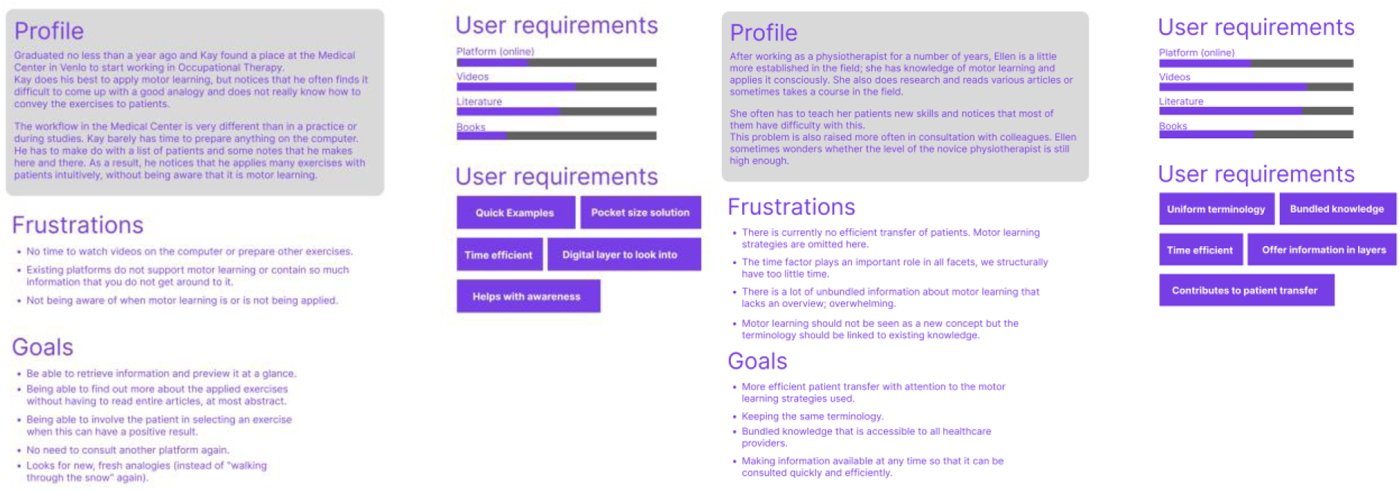
The fictional personae represented the less experienced (Kay Wolters, left) and experienced physiotherapist (Ellen Beckers, right). Their challenges (“frustrations”) and needs (“goals”) to make underpinned choices and support the application of motor learning strategies were described.

*Kay Wolters*, who represented the younger, less-experienced therapists, needed practical examples that were presented in an accessible way and raised his awareness with respect to motor learning, ideally in both an analog and digital way. *Ellen Beckers*, representing more-experienced therapists, needed uniformity in motor learning terminology, bundled knowledge that could be consumed in a time-efficient way, and in-depth information (e.g., scientific articles).

The creation of the two personae formed the basis for the follow-up session in which a list was created that specified the user requirements (see Appendix A). In short, the extension of the framework had to inspire, layer information (i.e., by having different sources of accessibility, from quick inspirational material to more in-depth information), save time, be suited to community-based physiotherapy practices, and have both an analog and a digital component. In the *ideate phase*, brainstorming sessions were held with the physiotherapists to generate ideas and possible solutions to tackle the challenges and needs specified in the previous phases. The “brainwriting” method was used; all therapists were asked to formulate ideas on note-taking sheets, which were then rotated clockwise.^24^ The therapists were then asked to add to the previous ideas without criticizing them; this process was repeated until saturation occurred. This working method was implemented to stimulate creativity.^25^ Finally, the different ideas, such as a mobile application, card deck, booklet, or online platform, were discussed with the group.

In the *prototype and test phases*, multiple ideas were developed as prototypes and presented to the therapists, after which feedback was received. In the first session, low-fidelity prototypes were shown to the group of physical therapists that were then discussed in a plenary session. During the discussion, several prototypes were discarded through communal voting. The remaining prototypes were further developed according to the input from the therapists and presented in a follow-up session. The researchers and designers had the final say on whether a prototype was selected for development.

### Final prototype of the extended framework

The final prototype consisted of a motor-learning card deck of 33 cards^a^ (*Fig. 4*), and website^b^ in which the cards were presented digitally (*Fig. 5*). The card deck provided a primary *practical layer* of information with examples from everyday practice. For each learning strategy described in the framework, multiple cards were developed for seven task categories (walking, transferring, activities of daily living (ADL), balancing, standing up and sitting, sports and hobbies, and arm–hand function).

**Figure 4:**
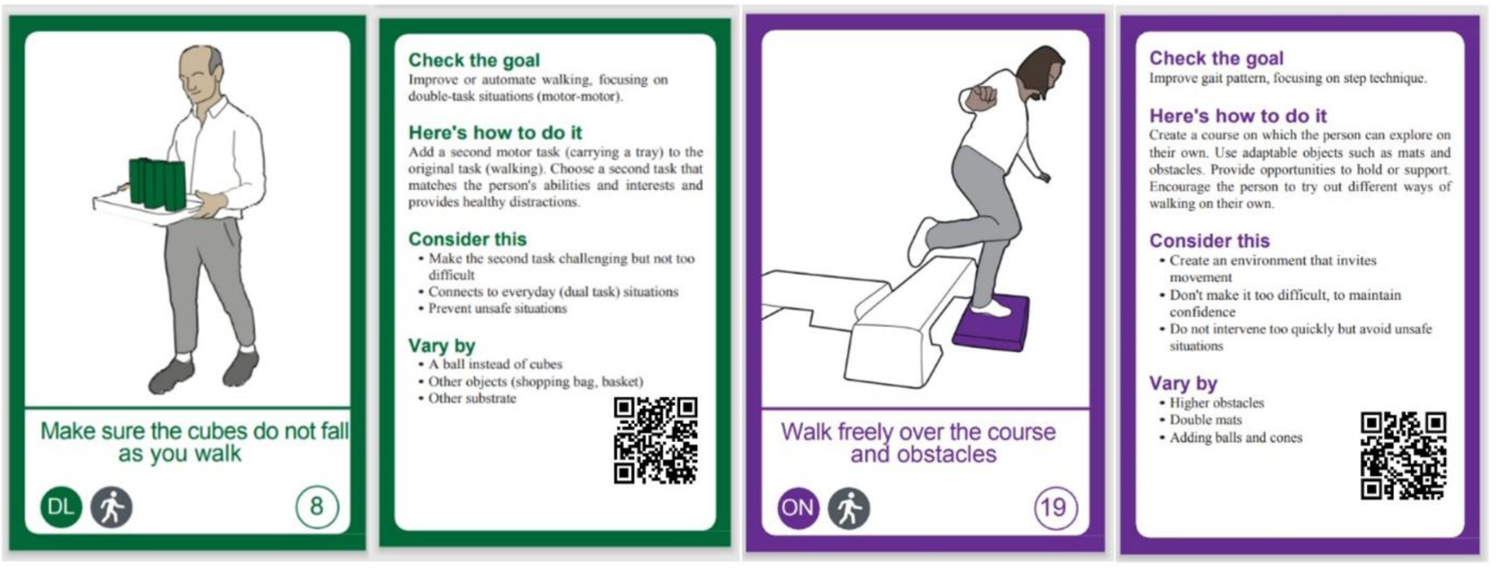
Examples of the developed cards. For each learning strategy described in the framework, multiple cards were developed for different tasks (e.g., walking, transfers, and activities of daily living). Each strategy has its own color, and each card is structured in the same way (Check the goal, Here’s how to do it, Consider this, and Vary by).

Additionally, on the website, a range of short theoretical (n = 8) and practical videos (n = 14) were produced that served as the secondary *sub-practical layer* for quick and accessible summaries and practical examples of the learning strategies. A tertiary *theoretical layer* of information was presented on the website describing the potential working mechanism and effects of the learning strategies with references to the most current scientific literature.

**Figure 5:**
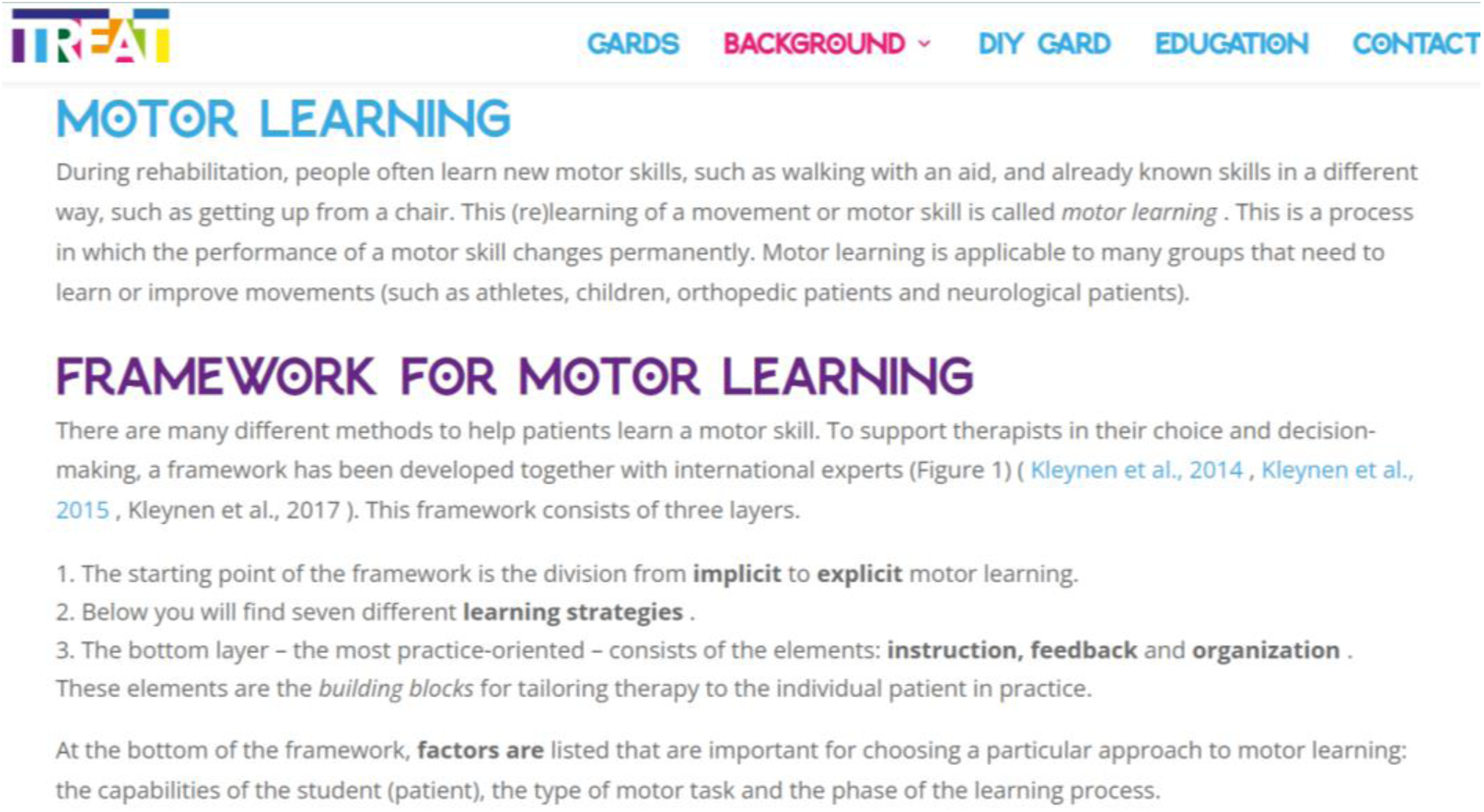
Screenshot from the motor-learning website showing a general page of background information regarding motor learning

It was deemed important to layer the information according to the needs of the different personae to avoid the risk of dissatisfaction.^26^ In addition, practical information was provided, such as which factors to consider when applying the strategy. Finally, the card deck and website were developed as an extension to the existing framework on motor learning (see Appendix B),^15^ hereafter referred to as the *extended framework*.

## Phase 2: Process evaluation of the extended framework

The process evaluation focused on how and why physiotherapists use and experience the extended motor-learning framework after six months of use.

## Methods

A process evaluation was conducted in which data on usage and experience were collected. Both ways of collection were deemed necessary to obtain the required information from the participants.^27,28^ The study was registered at ClinicalTrials.gov (NCT06390800).

### Participants

Thirteen therapists from the 10 included practices participated in the second phase of the study. Five therapists were already included in the user-centered design, and eight were newly recruited and thus unfamiliar with the content and results from the user-centered design. The therapists gathered information regarding their clinical decision-making with geriatric patients (65 years or older) with or without a neurological disorder who had at least one treatment goal related to improving motor skills. All participating therapists gave written informed consent.

### Preparatory training

To ensure that all therapists understood the extended framework and to increase compliance, a training session was provided. Two months prior to the training, all participating physiotherapists received the extended framework, which allowed them to familiarize themselves with the content. During this period, the physiotherapists were encouraged to ask questions if they encountered obstacles. The submitted questions were used to tailor the content of the preparatory training. To stimulate therapists’ compliance, every physiotherapist had to create a personal action plan regarding the use of the extended framework in their daily practice.

**Table 1:** Overview of outcome measures by objective of the process evaluation.

|  | Use of the extended framework | Experience with the extended framework |
| --- | --- | --- |
| Monitors (biweekly) | X |  |
| Digital questionnaires (T0, T1, and T2)* | X | X |
| Midterm evaluation (T1) |  | X |
| Final evaluation (T2) |  | X |

\* T0 = baseline, T1 = midterm, T2 = final

### Procedure

After completion of the preparatory training, the participating physiotherapists used the extended framework in their daily practice for six months. They were encouraged to use the extended framework but were free to decide how, and for which patients, as part of their usual care. The usage of and experience with the extended framework were assessed through several measures at baseline (T0), midterm at three months (T1), and at six months (T2) (see *Tab.1*).

### Outcome measures

#### Monitor

A biweekly monitor was developed to observe the frequency of using the extended framework, the patient population treated (multiple-choice questions), the learning strategies applied (multiple-choice questions), and the underlying reasons for applying this motor-learning strategy (free text box). Each participant could monitor the questions in their own Microsoft Word file.

#### Digital questionnaires

The self-developed digital questionnaire, using QuestBack (version 1.7, 2024, Oslo, Norway), was administered three times (T0, T1, and T2). Both open-ended (free text box) and closed-ended questions (11-point Likert scale) were used to measure the therapists’ experiences with the extended framework. Although some questions overlapped across timepoints, the focus was different. For T0, the emphasis was on therapists’ expectations, while, for T1 and T2, the focus was on their experiences with the extended framework, the perceived usefulness and ease of use of the extended framework, and self-efficacy regarding motor learning.

#### Evaluation sessions

A midterm and final evaluation were organized for all participating physiotherapists. During the midterm evaluation, a user journey matrix was created to identify and further specify user problems, brainstorm on solutions for these problems, and determine any necessary adjustments to the extended framework.^23^ For the final evaluation, the goal was to reach a definitive version of the extended framework based on the experiences from the therapists and communally brainstorm on the dissemination and valorization of the material. The session was used to determine which adjustments could and should be realized. The nominal group technique was used to ensure active participation from all participants and reach a consensus on the final list of adjustments.^29,30^ The collected data during the final evaluation were added to the existing user-journey matrix to ensure completeness.

### Data collection and analysis

The data from the monitors and digital questionnaires were analyzed using descriptive statistics, such as frequency tables, means and standard deviation (when parametric), or medians and interquartile range (when nonparametric). To visualize changes over time, Likert scale data from the digital questionnaires were visualized using boxplots. The answers provided to open-ended questions and feedback in textboxes from both the monitor and digital questionnaires were clustered into main and sub-themes by learning strategy in a Microsoft Excel file by one researcher, after which point the data were manually checked per cluster by a second researcher.

The data from the midterm and final evaluation were digitally transcribed verbatim into a Microsoft Word file, then manually checked and corrected. The transcriptions were then clustered into main and subthemes and processed into the *user journey*. The topics were divided into *feelings/intentions, gains, barriers*, *and opportunities/needs.* A concept version of the table was presented to the therapists for a member check. Quotations were used for illustration purposes.

## Results

At the commencement of the process evaluation, 13 physiotherapists were included. Between the T0 and T1 measurement of the study, three participants dropped out because of a lack of time, resulting in one community-based practice’s discontinuing its participation. The participant characteristics at the start of the study are displayed in *Tab. 2.* The data of 205 monitors (therapy sessions) from 80 different patients were collected. The digital questionnaires had a 100% response rate at each of the three time points (n = 13, n = 10, and n = 10, respectively). The midterm evaluation had an attendance rate of 50% (n = 5), and a final evaluation of 90% (n = 9).

**Table 2:** Demographics of the physiotherapists.

|  |  |
| --- | --- |
| Participants ($n$) | 13 |
| Women ($n$) | 9 |
| Age in years (range) | 35.8 (23–56) |
| Experience in years (range) | 12.5 (2–33) |
| Weekly patient contact hours (range) | 21.5 (12–35) |
| Degree of education ($n$) | Bachelor, 10 |
|  | Master, 3 |

### Usage of the extended framework

Based on data from the questionnaires, the use of the extended framework varied from regularly (T1 = 20%, T2 = 10%) to barely (T1 = 70%, T2 = 70%) to never (T1 = 10%, T2 = 20%). Commonly reported reasons for not using the extended framework included a lack of time and motivation, as well as the material’s not fitting into the physiotherapists’ daily routines, both in terms of clinical reasoning and actual practice.

The most represented diagnoses were Parkinson’s disease (32.5%) and stroke (32.5%) (*Tab. 3*). The most applied learning strategies were errorless learning (29%) and dual task learning (21%; *Tab. 4*).

In 165 therapy sessions, the participating physiotherapists used sources other than the extended framework to support their decision-making regarding motor learning (e.g., books, websites), earlier acquired knowledge and skills (e.g., courses and workshops), and their own expertise or experience. From the extended framework, the card deck was used most (43 sessions). Information and videos on the website were used in only 15 and four therapy sessions, respectively.

**Table 3:** Data of the therapy sessions that were monitored (biweekly).

|  | *Number of patients* | *(%)* |
| --- | --- | --- |
| *Age category* |  |  |
| 65–70 | 24 | 30 |
| 71–75 | 19 | 24 |
| 76–80 | 20 | 25 |
| 81–85 | 7 | 8.75 |
| 86+ | 10 | 12.5 |
| *Diagnosis* |  |  |
| Parkinson's disease | 26 | 32.5 |
| Dementia | 12 | 15 |
| Stroke | 26 | 32.5 |
| Other neurological diseases | 14 | 17.5 |
| General geriatric health problems | 8 | 10 |
| Other orthopedic health problems | 10 | 12.5 |

**Table 4:** Learning strategies used by therapist.

|  | *Number of sessions* | *(%)* |
| --- | --- | --- |
| *Applied learning strategy* |  |  |
| Analogy learning | 2 | 15.5 |
| Discovery learning | 56 | 12 |
| Trial and error | 41 | 8.75 |
| Errorless learning | 136 | 29 |
| Observational learning | 49 | 10.5 |
| Movement imagery | 15 | 3 |
| dual-task learning | 99 | 21 |
| Other strategy | 1 | 0.25 |

The data from the monitors that provided reasons for clinical decision-making were divided into 11 themes across the learning strategies (see *Appendix C*). For analogy learning, the most common reasons for application were that it best suited the activity or that there was a movement pattern to be improved.

> *‘‘I used the marching analogy to stimulate his gross motor skills and ultimately his gait.’’ (Physiotherapist 07)*

For errorless learning specifically but also for discovery learning, the most common reason for application was stimulating self-confidence regarding movement.

> *‘‘The patient often falls and gets more unsure over time; with this strategy, I hope to give him more confidence.’’ (Physiotherapist 03)*

The most common reason for applying dual-task learning was the patient’s struggle with dual tasks occurring in daily life.

> *‘‘He often needs to cross the street with his grandson, meaning he has to walk, watch out, and talk simultaneously.’’ (Physiotherapist 12)*

Therapists chose trial and error to challenge their patients and allow them to discover what worked and where their boundaries lay.

> *“Especially given the diversity of manifestations within neurological issues, I find it important that the movement solution is found by the patient themselves.’’ (Physiotherapist 14)*

In observational learning, the primary objectives for application were to reduce explicit instructions and uncover patients’ personal limitations.

> *‘‘I first showed the exercise myself, which is more clear than verbal explanations.’’ (Physiotherapist 03)*

For analogy learning, discovery learning, errorless learning, movement imagery, and dual-task learning, often no explanation was provided for why this strategy was chosen; rather, a general statement or explanation of the exercise was provided.

> *‘‘I let him walk in between two surfaces, whereafter I narrow the pathway and he can try again.’’ (Physiotherapist 10)*

### Experiences with the extended framework

The therapists rated the card deck with a median of 7 (ranging from 2 to 8) and the website with a median of 7.5 (ranging from 4 to 10).

Trends across time were observed in “Intention to use” (a negative trend over time) and “Not having to think much when using the extended framework” (a positive trend over time; *Fig. 6*). Moreover, a slight increase was observed in “having knowledge to apply the extended framework” and “confidence regarding motor learning.”

**Figure 6:**
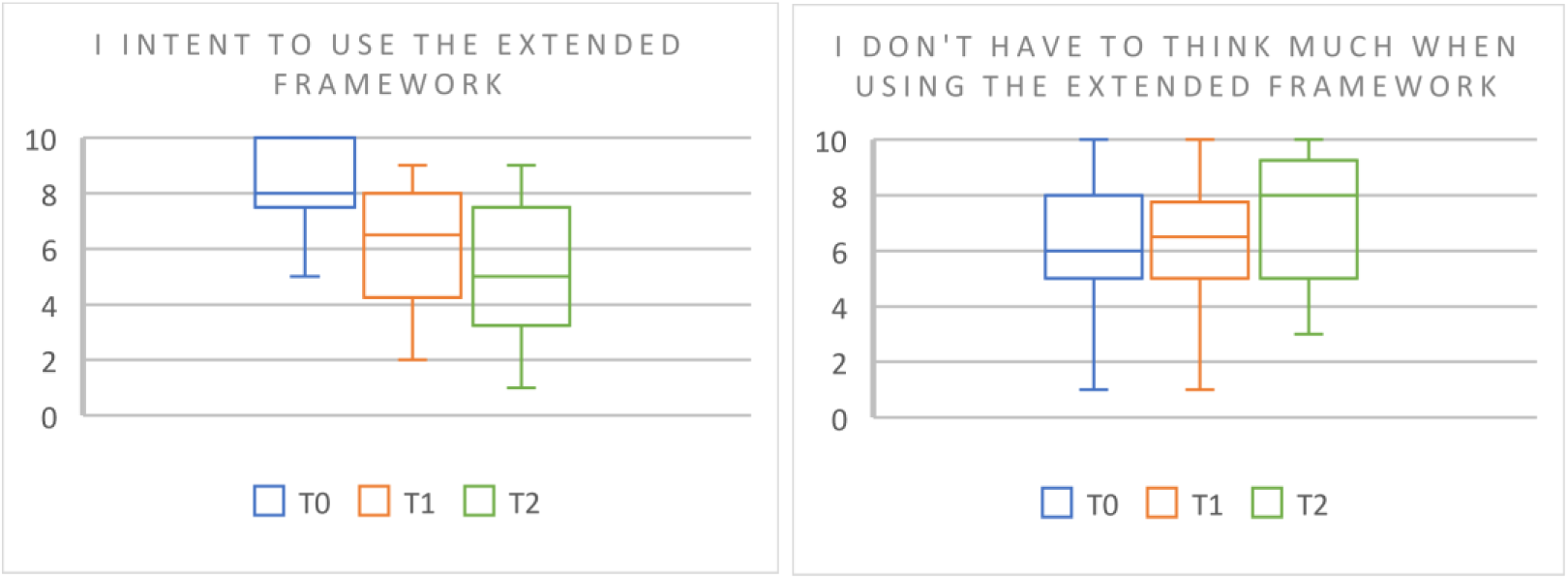
Boxplots from the digital questionnaires. Digital questionnaires: 0 completely disagree – 10 completely agree, T0 = baseline, T1 = midterm, T2 = final.

The results from the midterm and final evaluation were processed into a *user journey matrix* (*Appendix D*). The evaluations showed that the benefits of receiving the extended framework, according to the therapists, were “efficient knowledge transfer,” “more motivation from therapists and patients about motor learning,” and “stimulation to explore novel treatment options.” The most challenging part for therapists seemed to be making time to study the material and stepping out of their comfort zone when applying unfamiliar motor-learning strategies and breaking routines. The physiotherapists stated that the website had the most potential for future implementation considering its dynamic structure: i.e., the possibility to add more sources and examples. Furthermore, the physiotherapists concluded that the extended framework had great potential for educational use.

## Discussion

The objectives of this study were to determine which supplementary knowledge and tools were needed to make the existing framework more usable in clinical practice for community-based physiotherapists and gather information on how they used and experienced the framework.

### Development of the extended framework (user-centered design)

The different subphases of the user-centered design were completed successfully. There was diverse and rich input from the different physiotherapists to outline their challenges and needs to create a concrete list of requirements. Unexpectedly, there was little need for more knowledge but rather a need for structure in the currently available literature. The personae revealed a noticeable difference between the less-experienced and more-experienced physiotherapists. Whereas the former needed more guidance to get acquainted with motor learning, the latter sought more in-depth knowledge on how the strategies work and why. The extended framework tried to play into the different needs by layering the information from inspiring examples on the cards to explanation of theory, working mechanisms, and potential effects on the website.

### Use of and experiences with the extended framework

The general use of the extended framework was limited throughout the study. Although rated higher, the website was used less frequently than the card deck. This may be attributed to physiotherapists’ struggle to integrate digital tools into their routines.^31,32^ Noticeably, physiotherapists stated that they mainly used ‘‘other sources’’ to support their clinical decision-making. These sources included their own experience or expertise: e.g., from previous courses and workshops. It could be argued that it took too much effort to read the material, prepare novel exercises, and try out new strategies. Choosing a habituated task was likely less demanding than exploring new treatment options. Moreover, physiotherapists mentioned that it was challenging and uncomfortable to break with their routines, especially when they felt the patient lacked understanding of a certain learning strategy.

In general, the physiotherapists had mixed experiences regarding the extension of the framework. On the one hand, they stated that the extended framework raised awareness and knowledge of motor learning, stimulated the exploration of new motor-learning options, elevated confidence with respect to motor learning, and increased the motivation of both the physiotherapist and patient in motor learning. On the other hand, they expressed that it was time-and energy-consuming, the cards were a simplification of the complexity of daily practice, and experienced physiotherapists might not gain much theoretical and practical knowledge from the extension. Perhaps due to their active participation in the design process, the expectations raised above the outcome, which can occur in highly involved cocreators.^26^ When the increased expectations are not met by the product outcome, dissatisfaction takes place; namely, the developed extension did have significant benefits according to the physiotherapists but probably still required too much time and energy to use.

The motor-learning strategies of errorless learning, dual-task learning, and analogy learning were applied most frequently. In general, these strategies can be regarded as promoting more implicit forms of motor learning, which varies from earlier research in which the therapist mainly used more explicit forms of motor learning.^33^ In many cases, reasons for applying certain strategies were in line with scientific studies.^15,34,35^ Still, the therapists frequently provided no explanation for why they chose a certain motor-learning strategy. When reviewing the results from the user journey matrix, we see that physiotherapists simply *feel* that some strategies work better than others, which is currently not evident from the literature yet.^6^ Along the same lines, therapists expressed the need for an instrument that helps determine the preferred strategies per subpopulation. Although the current literature suggests that personalization is important for the success of motor learning,^11–14^ no clear conclusions on the effects across populations can be drawn.^6^ Until the state of research has evolved, physiotherapists’ most convenient strategy seems to be reading into the existing theories of motor learning and being aware of the different motor-learning possibilities, including considering their respective effects.

### Study limitations

The current study was conducted in collaboration with researchers, physiotherapists, patients, and designers. This collaboration was implemented to ensure satisfaction from different perspectives and increase the likelihood of a successful extension of the framework.^36^ A pitfall of such collaboration, however, is the different roles one must adopt, such as, in this case, that of both designer and physiotherapist. The initially formulated needs during the user-centered design, in particular, do not seem to correspond to the actual needs found during everyday practice. It was previously observed that a physiotherapist might be distracted from the actual needs presented during practice by being involved in the creation process.^37^ The included sample of physiotherapists in this study was relatively small but with broad variation in terms of age and working experience. The sample was, however, employed in a similar working environment (i.e., community-based physiotherapy practices) and was self-selected, meaning that all therapists applied to participate in the study and therefore might be part of the ‘‘early adopters’’ or even ‘‘innovators’’ group.^38^ This limits how generalizable the observed results can be to different clinical settings.

### Conclusions and future research

The results from this study indicate that the physiotherapists regarded the developed extension of the framework as beneficial for exploring new motor-learning options, increasing knowledge and awareness, elevating confidence in motor learning, and motivation of both the therapist and patient in motor learning. It seemed, however, that the time and energy costs on top of the disruption of breaking one’s own routines outweighed the benefits of using the extended framework in a community-based setting. The physiotherapist saw the future of the extended framework as most promising in educational settings.

Future research should aim to determine whether the extended framework applies similarly in different clinical settings, such as hospitals or rehabilitation centers. We also suggest an assessment of whether physiotherapists in different settings reason similarly regarding the application of motor-learning strategies in the attempt to personalize motor learning. More Insight into thinking patterns with respect to clinical decision-making in diverse settings will help us better understand the complex field of motor learning in clinical practice.

## Data Availability

All data produced in the present study are available upon reasonable request to the authors

## Abbreviations

ADL: activities of daily living

## Acknowledgements

This study was executed in collaboration with Regtop Fysiotherapie, ART-health Fysiotherapie, Van Hoof centrum voor Therapie & Gezondheid, Fysiotherapie Zesgehuchten, Fysiotherapie Veldhoven Zuid, Fysio van Hoof.

## Statement

During the preparation of this work, the authors used the assistance of an AI language model provided by OpenAI, known as ChatGPT, to refine writing. After using this tool, the author(s) thoroughly reviewed and edited the content as needed and take full responsibility for the accuracy and integrity of the public.

This study was funded by Regieorgaan SIA (Dutch Organization for Scientific Research Applied Research Fund) under grant number RAAK.PUB09.001. The funding source had no involvement in the execution of the study or the analysis of the data. The second part of this study, the process evaluation, was registered at ClinicalTrials.gov (NCT06390800).

The authors declare no conflict of interest.

## Suppliers

A: developed by Oburon Design.

B: developed by JW communicatie.

## Appendix A: List of requirements

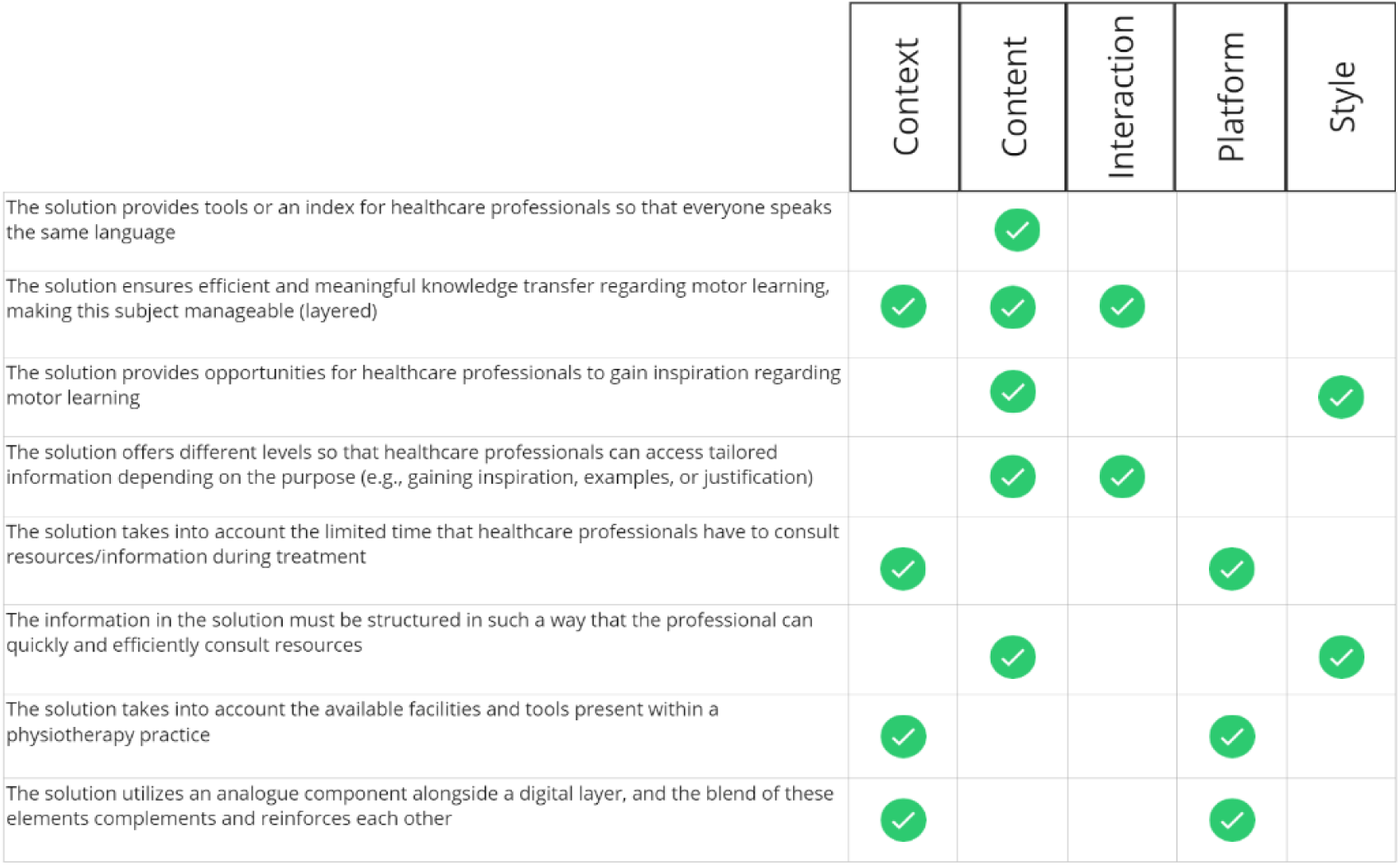

## Appendix B: Motor-learning framework 2020

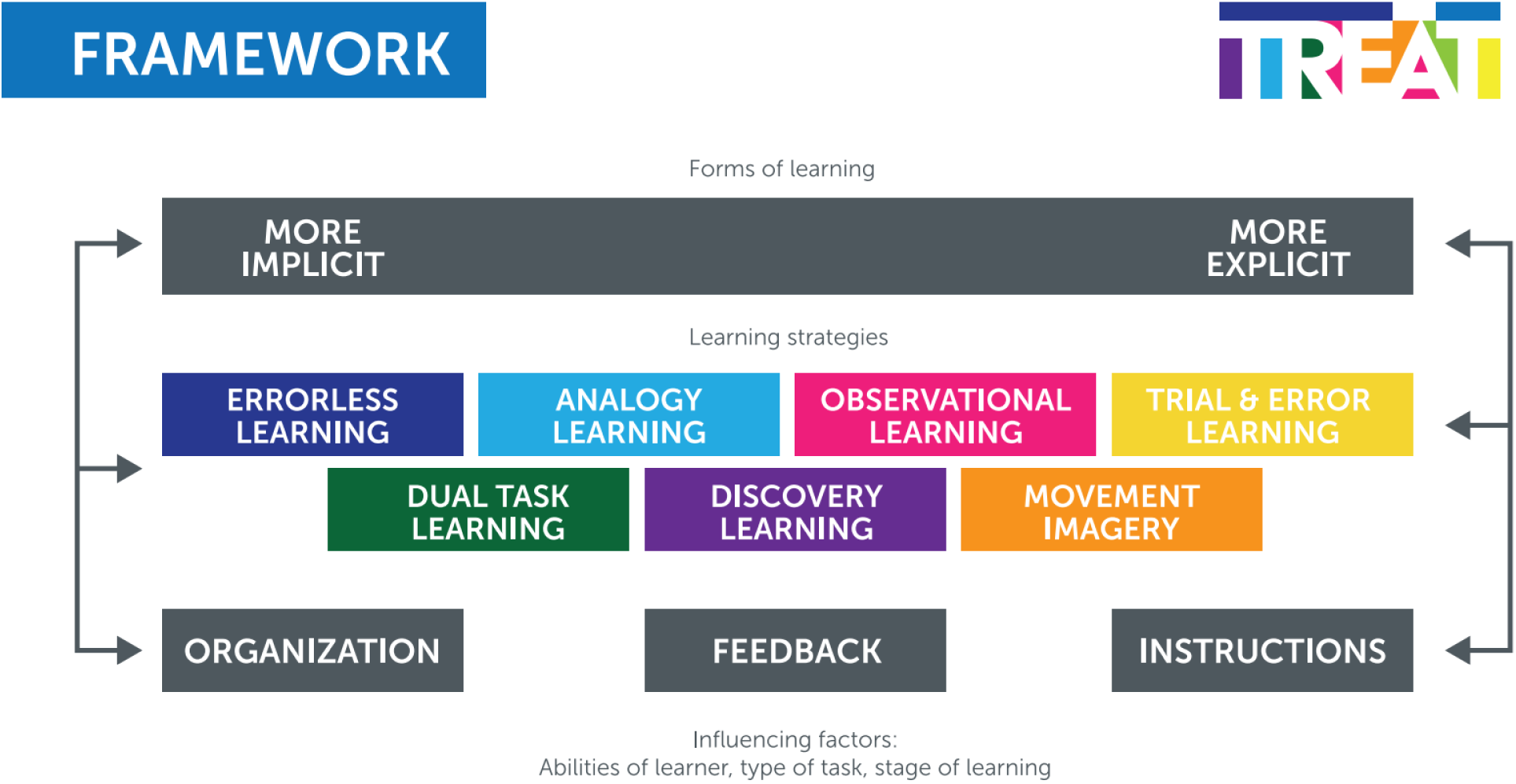

## Appendix 3: Goals by learning strategy

*Goals of therapists by learning strategy*
| *Main reason to choose a learning strategy* | *Analogy learning* | *Discovery learning* | *Trial & error* | *Errorless learning* | *Observational learning* | *Movement imagery* | *Dual-task learning* |
| --- | --- | --- | --- | --- | --- | --- | --- |
| To improve a specific motor skill | x | x |  | x |  | x | x |
| To take general cognitive problems into account | x |  |  | x |  |  | x |
| To circumvent explicit instruction | x |  |  | x | x |  |  |
| To increase self-confidence of the patient |  | x |  | x |  |  |  |
| To challenge the patient |  | x | x |  |  |  |  |
| To stimulate automatic movement | x |  |  | x |  |  | x |
| To explore new strategies as a therapist |  | x | x |  | x |  |  |
| To train dual tasks occurring in daily life |  |  |  |  |  |  | x |
| Example on card suited the activity best | x |  |  | x |  |  |  |
| Other strategies did not work |  |  |  |  |  | x |  |
| No specific explanation was provided | x | x |  | x |  | x | x |

*Most prevalent categories per learning strategy received a marking.*

## Appendix 4: User journey

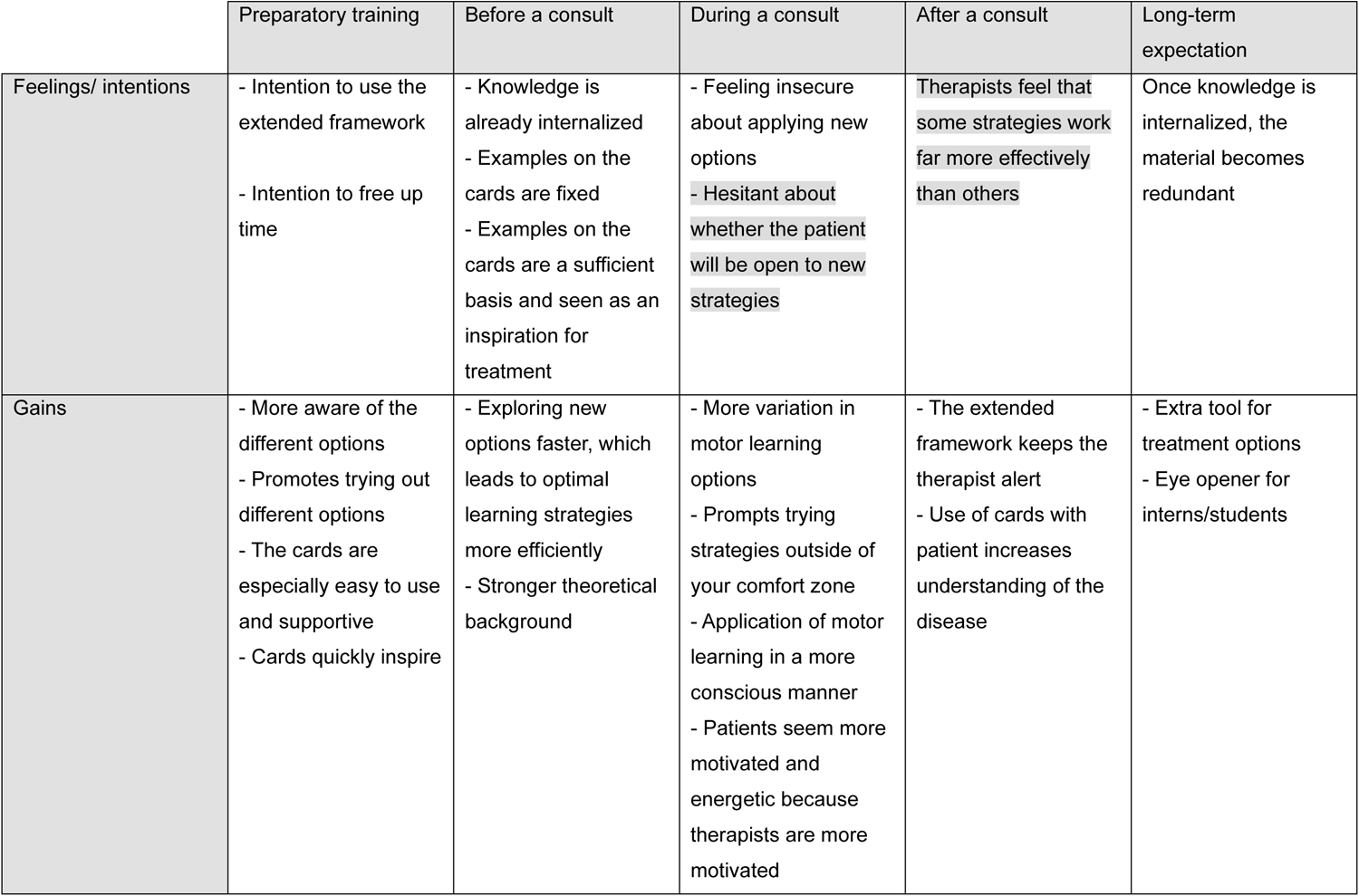
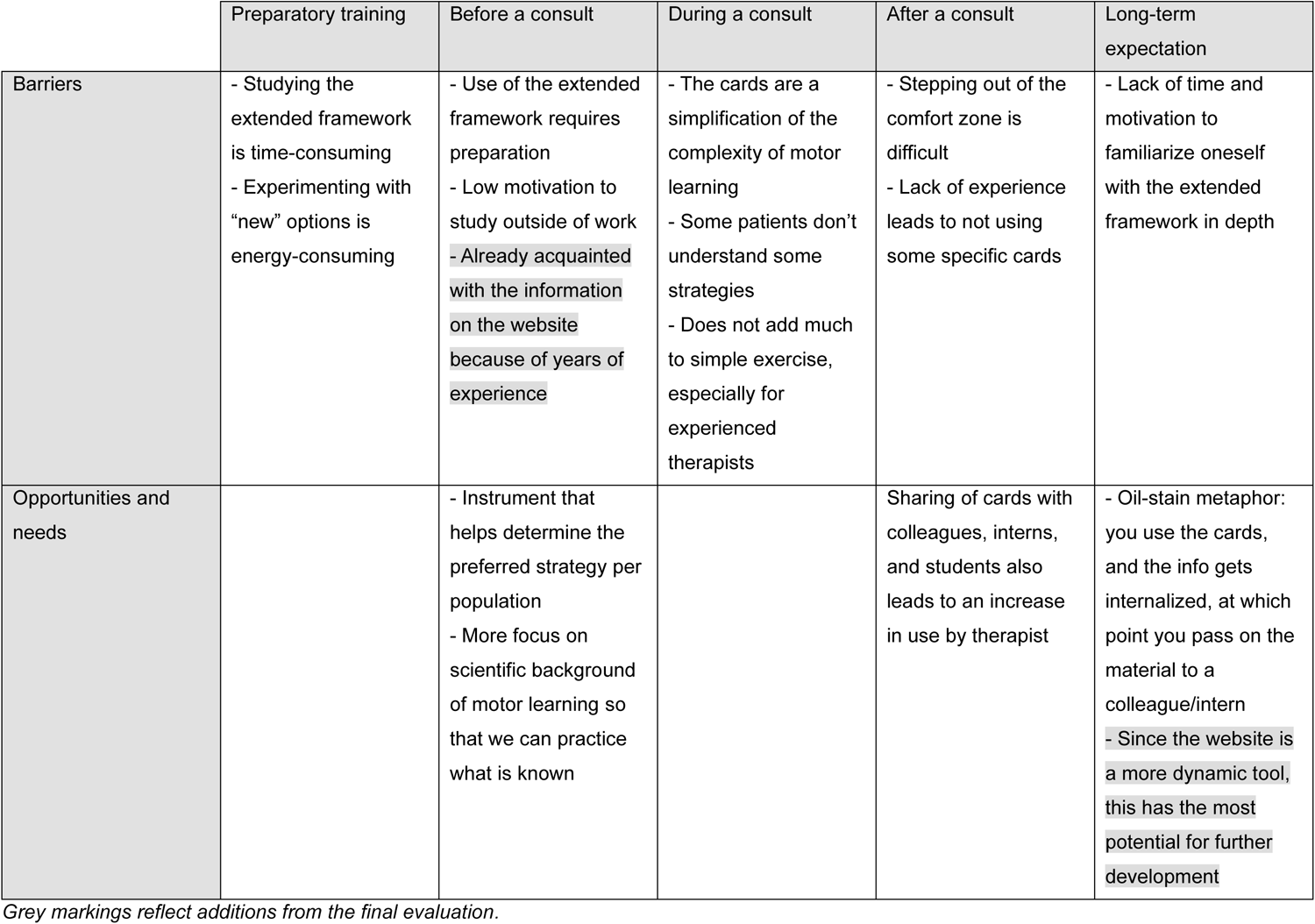

